# Common genetic variants associated with urinary phthalate levels in children: a genome-wide study

**DOI:** 10.1101/2024.02.02.24302120

**Authors:** Mariona Bustamante, Laura Balagué, Zsanett Buko, Amrit Kaur Sakhi, Maribel Casas, Lea Maitre, Sandra Andrusaityte, Regina Grazuleviciene, Kristine B Gützkow, Anne Lise Brantsæter, Barbara Heude, Claire Philippat, Leda Chatzi, Marina Vafeiadi, Tiffany C Yang, John Wright, Amy Hough, Carlos Ruiz-Arenas, Ramil Nurtdinov, Geòrgia Escaramís, Juan Ramon Gonzalez, Cathrine Thomsen, Martine Vrijheid

## Abstract

**Introduction:** Phthalates, or dieters of phthalic acid, are a ubiquitous type of plasticizer used in a variety of common consumer and industrial products. They act as endocrine disruptors and are associated with increased risk for several diseases. Once in the body, phthalates are metabolized through partially known mechanisms, involving phase I and phase II enzymes.

**Objective:** In this study we aimed to identify common single nucleotide polymorphisms (SNPs) and copy number variants (CNVs) associated with the metabolism of phthalate compounds in children through genome-wide association studies (GWAS).

**Methods:** The study used data from 1,044 children with European ancestry from the Human Early Life Exposome (HELIX) cohort. Ten phthalate metabolites were assessed in a two-void urine pool collected at the mean age of 8 years. Six ratios between secondary and primary phthalate metabolites were calculated. Genome-wide genotyping was done with the Infinium Global Screening Array (GSA) and imputation with the Haplotype Reference Consortium (HRC) panel. PennCNV was used to estimate copy number variants (CNVs) and CNVRanger to identify consensus regions. GWAS of SNPs and CNVs were conducted using PLINK and SNPassoc, respectively. Subsequently, functional annotation of suggestive SNPs (p-value <1E-05) was done with the FUMA web-tool.

**Results:** We identified four genome-wide significant (p-value <5E-08) loci at chromosome (chr) 3 (*FECHP1* for oxo-MiNP_oh-MiNP ratio), chr6 (*SLC17A1* for MECPP_MEHPP ratio), chr9 (*RAPGEF1* for MBzP), and chr10 (*CYP2C9* for MECPP_MEHPP ratio). Moreover, 113 additional loci were found at suggestive significance (p-value <1E-05). Two CNVs located at chr11 (*MRGPRX1* for oh-MiNP and *SLC35F2* for MEP) were also identified. Functional annotation pointed to genes involved in phase I and phase II detoxification, molecular transfer across membranes, and renal excretion.

**Conclusion:** Through genome-wide screenings we identified known and novel loci implicated in phthalate metabolism in children. Genes annotated to these loci participate in detoxification and renal excretion.

**Graphical abstract:** 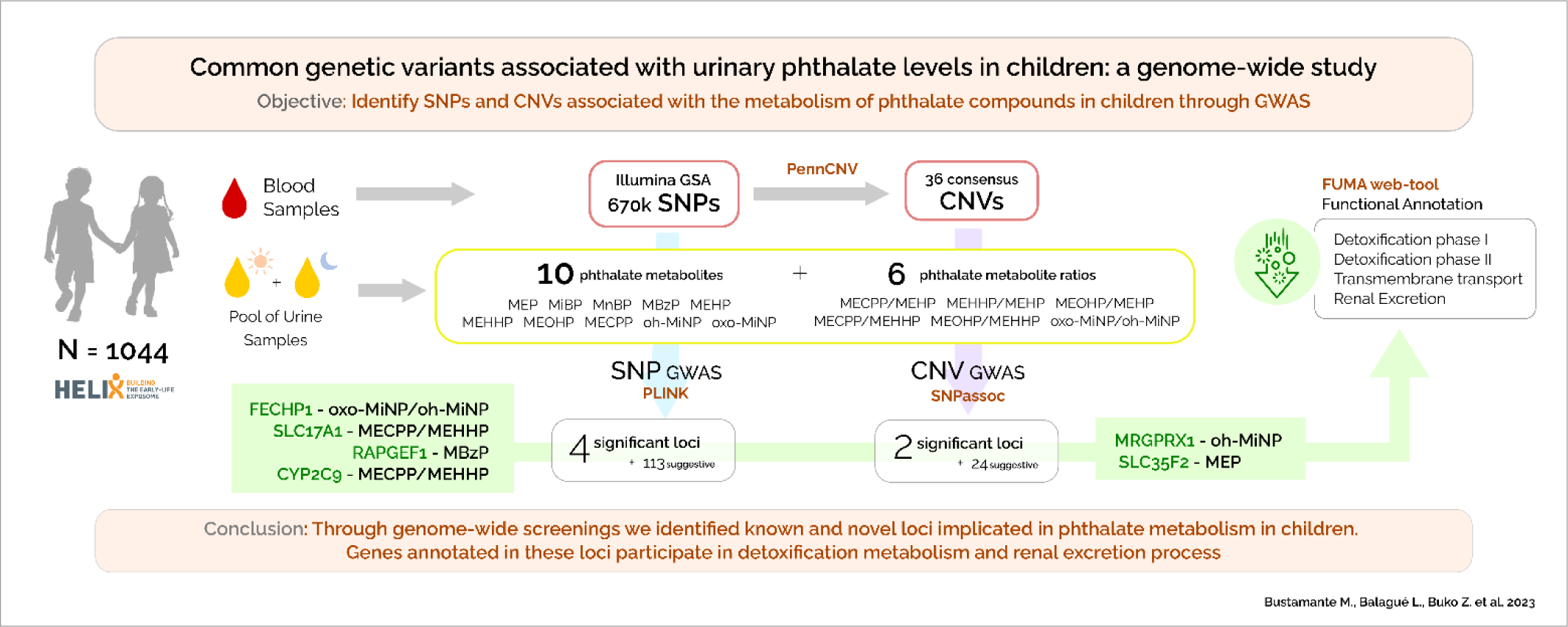

**Highlights:**

- The genetic variation involved in phthalate detoxification in humans is partially known.
- We identified four loci at genome-wide significance, and 113 at suggestive significance, some of them being novel.
- Two copy number variants were also identified.
- Functional annotation highlighted genes in phase I and II detoxification and renal excretion.

## 1. Introduction

Phthalates, or dieters of phthalic acid, are a ubiquitous type of plasticizer used in a variety of common consumer and industrial products, whose exposure is widespread and ever-changing due to our constantly evolving environment and habits (Praveena et al., 2018; Wang et al., 2019). Phthalates are known to be endocrine disrupting chemicals (EDCs), and epidemiological research has suggested that exposure to phthalates is associated with increased risk for several diseases including infertility, allergy, obesity, diabetes, and behavioral problems (Praveena et al., 2018; Wang et al., 2019). Since children are especially vulnerable to contaminants as they are still developing, there is a great concern over the potential for phthalate exposure to disturb normal growth and development (Braun, 2017; Casale and Rice, 2023; Lee et al., 2022, 2023).

Phthalates can be separated into low-molecular-weight (LMW, 3-6 carbon atoms) and high-molecular-weight (HMW, 7-13 carbon atoms) compounds (Praveena et al., 2018; Wang et al., 2019). LMW phthalates are used as solvents and are usually found in medications and personal care items such as deodorants, lotions, and shampoos. HMW phthalates are used in the manufacturing of flexible plastics for purpose of vinyl flooring, adhesives, medical devices, and food packaging. The main intake of LMW phthalates is through skin and inhalation, while HMW phthalates are incorporated into the body through ingestion (Kim and Park, 2014). In the European Union, some phthalates are now banned from use in cosmetics and regulated in material intended to come into contact with food.

After exposure, phthalates are rapidly metabolized and excreted mainly in urine as a result of phase I and phase II enzymes (Domínguez-Romero and Scheringer, 2019; Praveena et al., 2018). First, diester phthalates are hydrolyzed to monoester (primary metabolites) by phase I esterase and lipase enzymes (Bhattacharyya et al., 2022). Subsequently, LMW phthalates are primarily excreted in urine and feces as monoesters, without further metabolism. In contrast, HMW phthalates need further metabolism through hydroxylation and oxidation by phase I enzymes thus producing a number of oxidative metabolites (secondary metabolites). These oxidated metabolites can be directly excreted in the urine, or alternatively, can be additionally metabolized through phase II enzymes. Phase II detoxification consists of conjugation reactions where a compound is added to the parental compound to generate hydrophilic conjugates which can then be easily excreted in the urine. Phthalates and their metabolites can be measured in diverse biological specimens, such as blood, urine, breast milk, and feces. However, urinary phthalate metabolites are the most frequently used biomarkers to track exposure to phthalates (Sakhi et al., 2017; Wang et al., 2019).

Phase I and phase II enzymes are highly polymorphic in humans, with slow and fast metabolic phenotypes determined by single nucleotide polymorphisms (SNPs) and copy number variants (CNVs) (Pinto and Eileen Dolan, 2011). This differential detoxification capacity might modify phthalate effects in the body and is rarely considered in epidemiological studies. In vitro studies demonstrated the importance for phthalate metabolism of genetic polymorphisms in *Cytochrome P450 (CYP) monooxygenases*, a family of phase I detoxification enzymes (Choi et al., 2012).

Moreover, in humans, urinary phthalate metabolite levels were reported to be associated with genetic variants in *CYP monooxygenases* (*CYP2C9* and *CYP2C19)* and in *UDP-glucuronosyltransferases (UGT1A7)* in young adults (Stajnko et al., 2022), and with variants in *GSTP1* and *SOD2* in children (Wang and Karmaus, 2017). However, these studies were based on a priori knowledge of candidate genes and SNPs, likely missing additional loci important for phthalate metabolism. Genome-wide association studies (GWAS), which allow the interrogation of millions of common genetic variants, both SNPs and CNVs, can help to overcome this limitation and reveal new phthalate detoxification pathways (Visscher et al., 2017).

Here, we aimed to identify genetic variants related to phthalate metabolism in children. For this, we analyzed the association of genome-wide SNPs and CNVs with urinary levels of ten phthalate metabolites and six phthalate ratios in 1,044 European ancestry children from the Human Early Life Exposome (HELIX) project and performed functional annotation of the identified variants.

## 2. Materials and methods

### 2.1. Study population

HELIX project is an ongoing population-based birth control study in six birth cohorts from different European countries: (1) EDEN - Étude des Déterminants pré et postnatals du développement et de la santé de l’enfant, France; (2) Rhea - the Rhea Mother-Child Study in Crete, Greece; (3) KANC - Kaunus Cohort, Lithuania; (4) MoBa – the Norwegian Mother, Father and Child Cohort Study, Norway; (5) INMA - - INfancia y Medio Ambiente, Spain; (6) BiB - Born in Bradford, United Kingdom (UK) (Maitre et al., 2018). The HELIX project aims to implement novel exposure assessment and biomarker methods to measure the early-life exposure to multiple environmental factors and associate these with omics biomarkers and child health outcomes, thus characterizing the “early-life exposome”. The entire study population is 31,472 mother-child pairs. The subcohort study includes 1,304 children with exposure, phenotypes, and molecular data measured at age of 6-12 years. The current study selected 1,044 children of European ancestry with genome-wide genetic and phthalate metabolite levels available from the HELIX subcohort (Appendix A - Supplementary Figure 1). Child’s ancestry was predicted from genome-wide genetic data. The following variables were used to describe the population: child’s sex (male/female), child’s age (in years), child’s obesity in 3 categories (normal, overweight, obese) based on the World Health Organization (WHO) body mass index (BMI) classification, and self-reported maternal education (primary, secondary and university or higher).

### 2.2. Ethics approval and consent to participate

All studies were approved by the national research ethics committees and informed consent to participate was obtained for all participants.

### 2.3. Phthalate biomarkers measurement

Metabolites for 6 different phthalates were measured in a pool of two urine samples collected in the morning and at bedtime at the mean age of 8 years (Figure 1). An aliquot of the urine pool was analyzed for the determination of four LMW primary phthalate metabolites [monoethyl phthalate (MEP), mono-n-butyl phthalate (MnBP), mono-isobutyl phthalate (MiBP), mono benzyl phthalate (MBzP)], one HMW primary phthalate metabolite [mono-2-ethylhexyl phthalate (MEHP)], and five HMW secondary metabolites [mono-2-ethyl-5-hydroxyphenyl phthalate (MEHHP), mono-2-ethyl-5-oxohexyl phthalate (MEOHP), mono-2-ethyl-5-oxyhexyl phthalate (MECPP), mono-4-methyl-7-hydroxyoctyl phthalate (oh-MiNP), and mono-4-methyl-7-oxooctyl phthalate (oxo-MiNP)], using high performance liquid chromatography coupled to mass spectrometry (Haug et al., 2018). Both external control samples, in-house control samples and blank samples were analyzed in each batch of about fifty samples. The limits of detection (LOD) of the method ranged from 0.067 to 0.67 ng/mL. All the samples had values >LOD. MEHP, MEHHP and MEOHP had 34, 3 and 1 missing values, due to underperformance of sample quantification.

**Figure 1.**
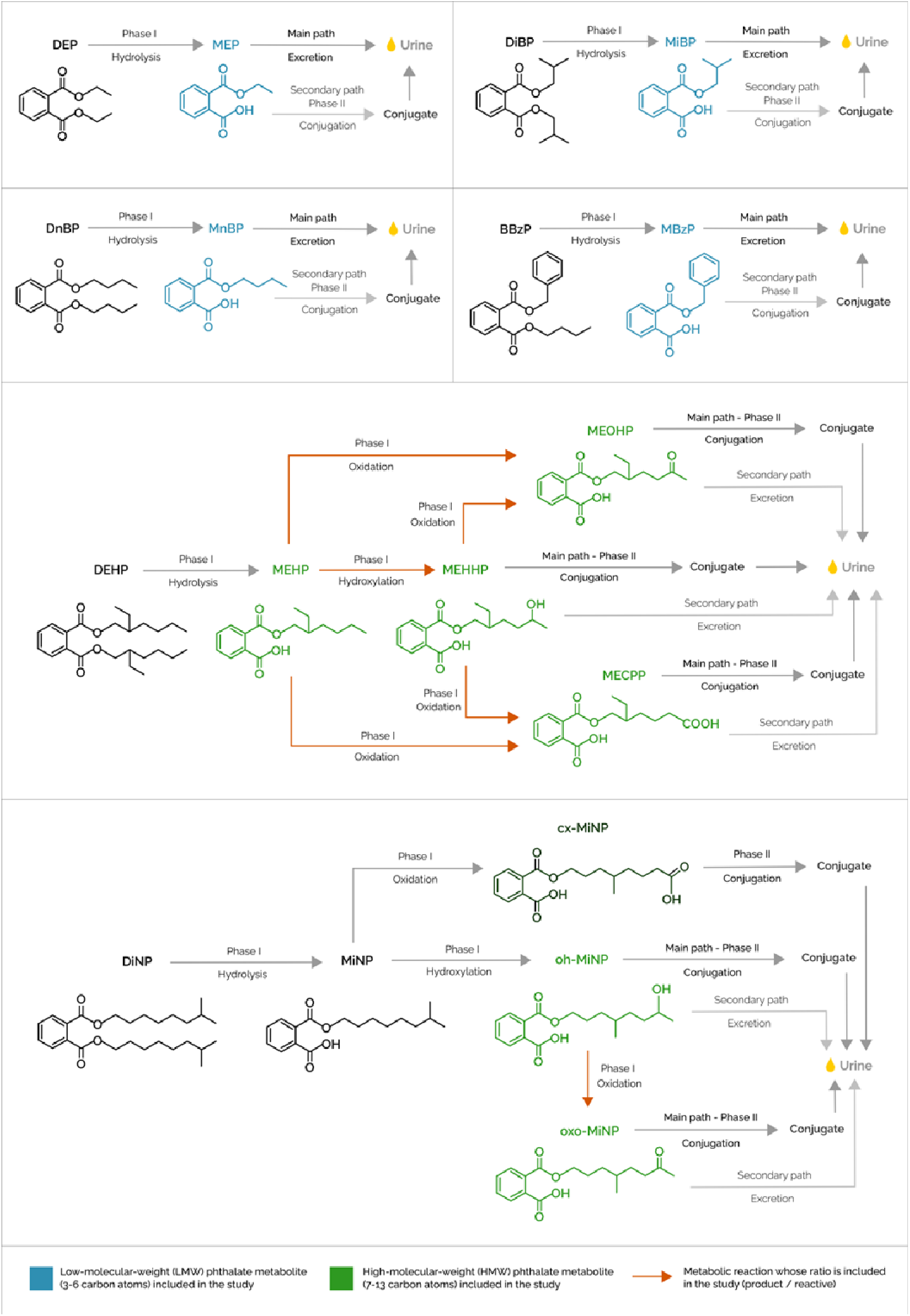
Summary of the phthalate metabolites and ratios included in the study. Diagram showing the phthalate metabolites included in the study, along with their precursor phthalates and intermediate metabolites. Low-molecular-weight metabolites (LWM) in the study are coloured in blue and high-molecular weight metabolites (HMW) in the study are coloured in green. Orange arrows represent the metabolic reactions whose ratio is included in the study (product/substrate).

To account for urine dilution, a second aliquot of urine was analyzed for creatinine concentration, and phthalate metabolite concentrations were divided by urinary creatinine levels (Haug et al., 2018). All creatinine-adjusted concentrations were log2 transformed to obtain normal distributions, as the original distributions were right-skewed. Finally, as a proxy of enzymatic activity, we calculated six ratios between product and substrate, in our case, between secondary and primary phthalate metabolites or between two secondary phthalate metabolites, as follows: MECPP_MEHP, MEHHP_MEHP, MEOHP_MEHP, MECPP_MEHHP, MEOHP_MEHHP and oxo-MiNP_oh-MiNP. The ratios were calculated using untransformed phthalate values unadjusted for creatinine, and then they were log2 transformed. Urine dilution was not taken into account as the ratios were computed using two compounds within the same urine sample.

### 2.4. Genetic data

DNA was obtained from buffy coats collected in EDTA tubes at the mean age of 8 years. DNA was extracted by cohort using a Chemagen kit in batched of 12 samples. Two techniques were used to determine the DNA concentration 1) NanoDrop 1000 UV-Vis Spectrophotometer (ThermoScientific) and 2) Quant-iT™ PicoGreen® dsDNA Assay Kit (Life Technologies).

Infinium Global Screening Array (GSA) MD version 1 (Illumina) was used for genome-wide genotyping at the Human Genomics Facility (HuGe-F), Erasmus MC (www.glimdna.org). Genotype calling and annotation were done using the GenTrain2.8 algorithm based on a custom cluster file implemented in the GenomeStudio software and the GSAMD-24v1-0_20011747-A4 manifest. SNP coordinates were reported on human reference GRCh37 and on the source strand.

PLINK program was used for the quality control of the genetic data (Purcell et al., 2007). Briefly, samples were filtered out if they had a call rate <97%, had sex inconsistencies, the heterozygosity was >3 standard deviations, if they were related (sharing >18.5% of alleles) or duplicated. For ancestry prediction from GWAS data, we used the Peddy program (Pedersen and Quinlan, 2017). Then genetic ancestry was contrasted with self-reported ethnicity and discordant samples were excluded. Genetic variants were filtered out if they had a call rate <95%, if they were in the non-canonical pseudo-autosomal region (PAR), if they had a minor allele frequency (MAF) <1%, and if they were not in Hardy-Weinberg equilibrium (HWE) at a p-value <1E -06. PLINK was also used to compute the first 20 principal components from the GWAS data of European ancestry children using the linkage disequilibrium (LD) clumping option. Ten PCs explained up to 19.2% of the genetic variance and the five first ones correlated with cohort.

Genome-wide imputation was performed with the Imputation Michigan server using the Haplotype Reference Consortium (HRC) panel, Version r1.1 2016. Before the imputation PLINK data was converted into VCF format and the variants were aligned with the reference genome. Eagle v2.4 was used for the phasing of the haplotypes and minimac4 for the imputation. In the end, we retrieved 40,405,505 variants after imputation. Then we filtered out the genetic variants according to the following parameters: imputation accuracy (R2) <0.9, MAF<5%, and HWE p-value <1E-06. After this the post-imputation dataset had 4,614,947 variants.

### 2.5. GWAS of SNPs

GWAS of SNPs were conducted with the PLINK program by applying a linear regression model for each phthalate metabolite and each SNP from chromosome 1 to 22, adjusting for sex, age and ten GWAS principal components as a proxy of ancestry (PCs) (Purcell et al., 2007). We run a total of 16 GWAS (five primary metabolites, five secondary metabolites, and six ratios). Genome-wide statistical significance was established at a p-value <5E-08, and suggestive statistical significance at a p-value <1E-5. The lambda genomic inflation factor was estimated as the median of the resulting chi-squared test statistics divided by the expected median of the chi-squared distribution. Quantile-quantile (QQ) plots were conducted with the ggplot2 package and Manhattan plots were conducted with the qqman package in R (D. Turner, 2018; Wickham, 2016). The lead SNP is defined as the SNP with the lowest p-value in a locus, and a locus is defined as a region of 1Mb.

In order to filter out associations between the SNPs and creatinine levels, we run sensitivity models adjusting for log2 transformed creatinine levels, instead of controlling (dividing) the phthalate concentrations by creatinine. Finally, we also run a GWAS for log2 transformed creatinine levels (g per L of urine).

### 2.6. GWAS of CNVs

Copy number variant (CNV) calls were obtained with PennCNV using BAF/LRR signal files from Illumina BeadStudio (Wang et al., 2007). A total of 22 samples from the initial 1,044 had to be removed due to poor quality in BAF or LRR values. After that, CNV calls overlapping with the centromeres were filtered out. After the quality control, consensus CNV regions were obtained with the CNVRanger R package following the reciprocal overlap procedure, that merges calls with sufficient mutual overlap, in our case 50% (Conrad et al., 2010; Da Silva et al., 2020). From the resulting consensus regions, only common CNVs with an alternative minor allele frequency (either gain or loss) >3% were kept for the analysis. Consensus CNV regions were classified into: (i) only loss, when we only detected deletions (CN=0 and CN=1); (ii) only gain, when we only detected duplications (CN=3; note that more than 3 copies were never detected); and (iii) loss and gain, when we detected both deletions and duplications (CN=0, CN=1 and CN=3).

Association analyses between each of the resulting regions and each of the 16 metabolites and ratios were performed with SNPassoc R package (González et al., 2007). Distinct association models were applied to each region based on the prior CNV classification. For only-loss regions we tested loss-codominant (0 vs 1 vs 2), loss-dominant (0+1 vs 2), loss-recessive (0 vs 1+2), loss-overdominant (0+2 vs 1) and loss-log-additive models; and for only-gain regions we tested gain-codominant (2 vs 3) model. Finally, for loss-and-gain regions we tested all loss-specific and gain-specific models above, as well as an additive lineal model taking into account loss and gain. All analyses were adjusted for the same covariates as the GWAS of SNPs (sex, age and ten GWAS principal components (PCs)). Multiple-testing correction was conducted by dividing the nominal p-value of 0.05 by the number of detected consensus CNVs (0.05/36 = 0.0014). As was done with SNPs, sensitivity models adjusting for log2 transformed creatinine levels were investigated in order to filter out associations between the CNVs and creatinine levels. Finally, we also run a model for log2 transformed creatinine levels (g per L of urine).

Annotation of genes in each CNV region was done with the biomaRt R package (Durinck et al., 2009).

### 2.7. Functional annotation of genetic variants, gene-wide association analysis and enrichment

For the annotation, prioritization, and biological interpretation of the genetic variants identified in the GWAS, we used the Functional Mapping and Annotation of Genome-Wide Association Studies (FUMA) tool (Watanabe et al., 2017). The SNP2GENE module computes LD structure, annotates SNPs using information from different databases, and prioritizes candidate genes. FUMA was run using default parameters except that the p-value threshold was set to 1E-05 instead of 1E-08. For the SNP annotation we used: ANNOVAR, CADD, RegulomeDB, expression quantitative trait locus (eQTL) mapping based on Genotype-Tissue Expression (GTEx) v8 and other databases, comparison with SNPs reported in the GWAS catalog and gene mapping through chromatin interaction maps. Then, with the GENE2FUNC module from FUMA prioritized genes from SNP2GENE module were used to identify shared molecular functions using different databases: Gene Ontology (GO) terms, Kyoto Encyclopedia of Genes and Genomes (KEGG), and Reactome, among others. GENE2FUNC was also used to assess enrichment for 54 tissues included in the GTEXv8 dataset.

For gene mapping through chromatin interaction we also used the EPIraction tool (Nurtdinov and Guigó, n.d.). Briefly, the tool contains a catalogue of tissue specific gene-enhancer interactions identified by applying linear models between gene expression and activities of nearby enhancers across 1,529 samples from 77 different tissues. In this study, we annotated SNPs to enhancers and then linked enhancers to genes in two candidate tissues for phthalate detoxification: liver and kidney.

## 3. Results

### 3.1. Description of the population

The description of the 1,044 HELIX participants is shown in Table 1. All participants were of European ancestry and from six different countries: Greece, Norway, Spain, Lithuania, France, and the UK. Forty-six percent of the participants were female, and the mean age of phthalate measurements was 8 years (standard deviation (SD): 1.6). Around half of the children were born from highly educated mothers. Urinary phthalate levels (micrograms (μg) per gram (g) of creatinine) were in the range or higher than levels reported in children and adolescents from 12 European countries for the period 2014–2021 (Vogel et al., 2023). After log2 transformation, levels tended to be normally distributed and the correlation among them ranged from 0.18 to 0.98 (Appendix A – Supplementary Figure 2). Highest correlations (r>0.79) were observed among MECPP, MEHHP, MEOHP, MEHP, which are all primary or secondary metabolites of the same parental compound, DEHP. The correlation of oh-MiNP and oxo-MiNP was also high (R=0.73).

**Table 1.** Descriptive of the study population. For continuous variables mean and SD are shown, while for categorical variables the sample size and its percentage % are reported. LMW: low molecular weight. HMW: high molecular weight. N=1,044.

| Variable | N (%), Mean (SD) | N missing |
| --- | --- | --- |
| Cohort |  |  |
| BiB (UK) | 90 (8.62%) | 0 |
| EDEN (France) | 135 (12.93%) | 0 |
| KAUNAS (Lithuania) | 196 (18.77%) | 0 |
| MoBa (Norway) | 239 (22.89%) | 0 |
| RHEA (Greece) | 186 (17.82%) | 0 |
| INMA (Spain) | 198 (18.97%) | 0 |
| Child's sex |  |  |
| Females | 473 (45.31%) | 0 |
| Males | 571 (54.69%) | 0 |
| Child's age (years) | 7.96 (1.54) | 0 |
| Child's obesity |  |  |
| Normal | 830 (79.50%) | 0 |
| Overweight | 155 (14.85%) | 0 |
| Obese | 59 (5.66%) | 0 |
| Maternal education |  |  |
| Primary | 115 (11.02%) | 0 |
| Secondary | 353 (33.81%) | 0 |
| University or higher | 550 (52.68%) | 0 |
| Urinary creatinine levels (g per ml of urine) | 1.004 (0.350) | 0 |
| Urinary LMW phthalate levels (µg per g of creatinine) |  |  |
| MBzP | 8.85 (16.14) | 0 |
| MEP | 77.32 (184.98) | 0 |
| MiBP | 57.48 (60.29) | 0 |
| MnBP | 33.78 (37.75) | 0 |
| Urinary HMW phthalate levels (µg per g of creatinine) |  |  |
| MEHP | 4.67 (11.09) | 34 |
| MECPP* | 53.56 (131.78) | 0 |
| MEHHP* | 31.20 (84.53) | 3 |
| MEOHP* | 18.80 (46.17) | 1 |
| oh-MiNP | 9.09 (16.20) | 0 |
| oxo-MiNP** | 5.62 (15.82) | 0 |

\*Metabolites of MEHP, \*\*Metabolite of oh-MiNP

### 3.2. GWAS of SNPs

To identify SNPs associated with the detoxification metabolism of phthalates in children, we run linear regressions for each of the ten phthalate metabolites and the six metabolic ratios adjusting for sex, age, and 10 GWAS principal components.

Genomic inflation factors (lambdas) ranged from 0.989 to 1.021 (Appendix A – Supplementary Figure 3). Four loci reached genome-wide significance (p-value <5E-08) as shown in the Miami plot (Figure 2). They involved two SNPs at chr3 associated with the oxo-MiNP_oh-MiNP ratio (lead SNP rs80064213_C, effect = 0.331, p-value = 4.15E-08, near *FECHP1* gene), 116 SNPs at chr6 associated with the MECPP_MEHPP ratio (rs1359232_A, effect = -0.134, p-value = 4.58E-17, *SLC17A1*), one SNP at chr9 associated with MBzP (rs138702233_T, effect = 0.394, p-value = 4.29E-08, *RAPGEF1*), and 658, 186 and 369 SNPs at chr10 associated with the MECPP_MEHPP, MEOHP_MEHHP and MECPP_MEHP ratios, respectively (rs74494115_T, effect = -0.309, p-value = 1.45E-63, *CYP2C9*). The locus zoom plots of these four loci are shown in Figure 3.

**Figure 2.**
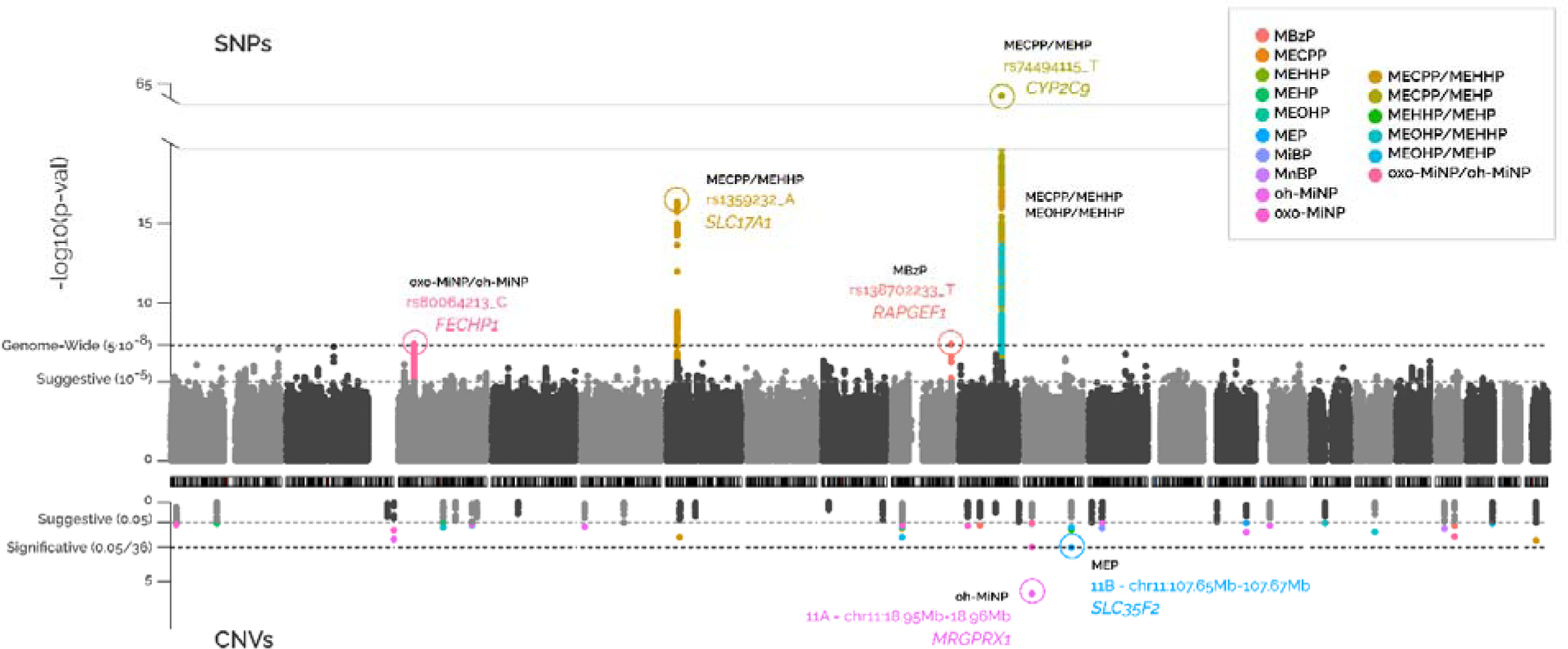
Miami plot of the association between common SNPs (top panel) and CNVs (bottom panel) vs phthalate levels or ratios. Each dot represents the association of a SNP or CNV. The x-axis indicates the position of the SNP or CNV in the genome. The y-axis shows the statistical significance, the –log10(p-value). Colours indicate the phthalate compoun or ratio the SNPs or CNVs are associated with. The SNPs/CNVs passing multiple-testing correction (5E-08 for SNPs and 1.39E-03 for CNVs) are annotated to the closest gene.

**Figure 3.**
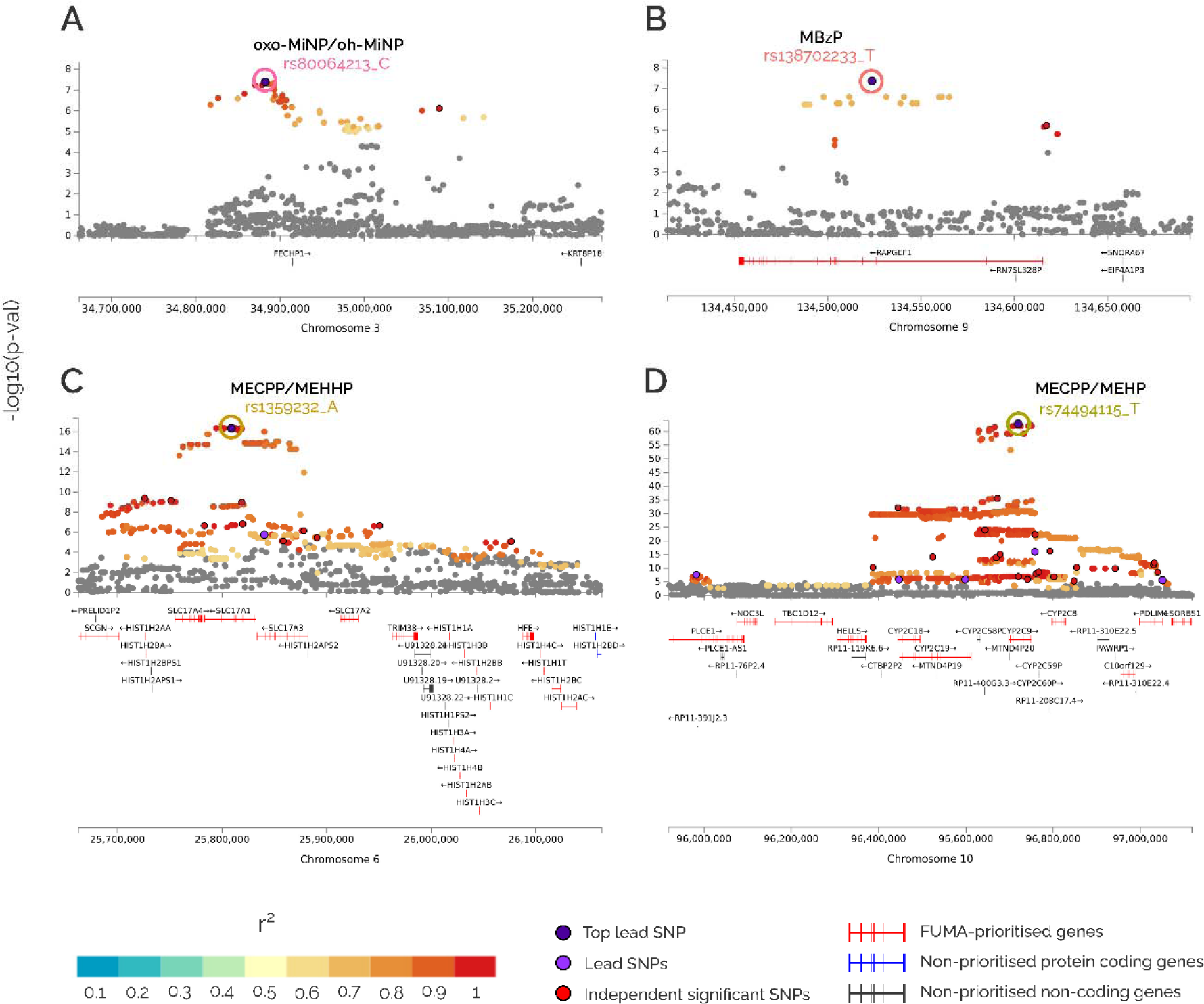
Locus zoom of the SNP-associated regions identified after multiple-testing correction. Each dot represents the association of a SNP. The lead SNPs are coloured in dark purple, other lead SNPs (SNPs i the region with LD <0.1 with top lead SNP) in light purple and independent significant SNPs in red. Other SNPs are coloured according to their linkage disequilibrium (r^2^) with the lead SNP. The y-axis shows the statistical significance, the –log10(p-value). The x-axis indicates the position of the SNP in the genome. FUMA prioritize coding genes and FUMA non-prioritized coding and non-coding genes are shown.

Moreover, 2,140 SNPs representing 158 loci (113 unique) were identified at suggestive significance (p-value <1E-05) for at least one phthalate metabolite or ratio (Table 2, Appendix B - Supplementary Table B1). The number of loci with suggestive statistical significance per phthalate metabolite or ratio was between five and 15. Thirty of the 117 suggestive or genome-wide significant unique loci were shared between more than one phthalate compound or ratio (Appendix A – Supplementary Figure 4). The genome-wide significant loci at chr3 (*FECHP1)*, chr6 (*SLC17A1)* and chr10 (*CYP2C9)*, also had suggestive associations with other phthalate compounds or ratios (Table 3, Appendix B – Supplementary Table B1).

**Table 2.**
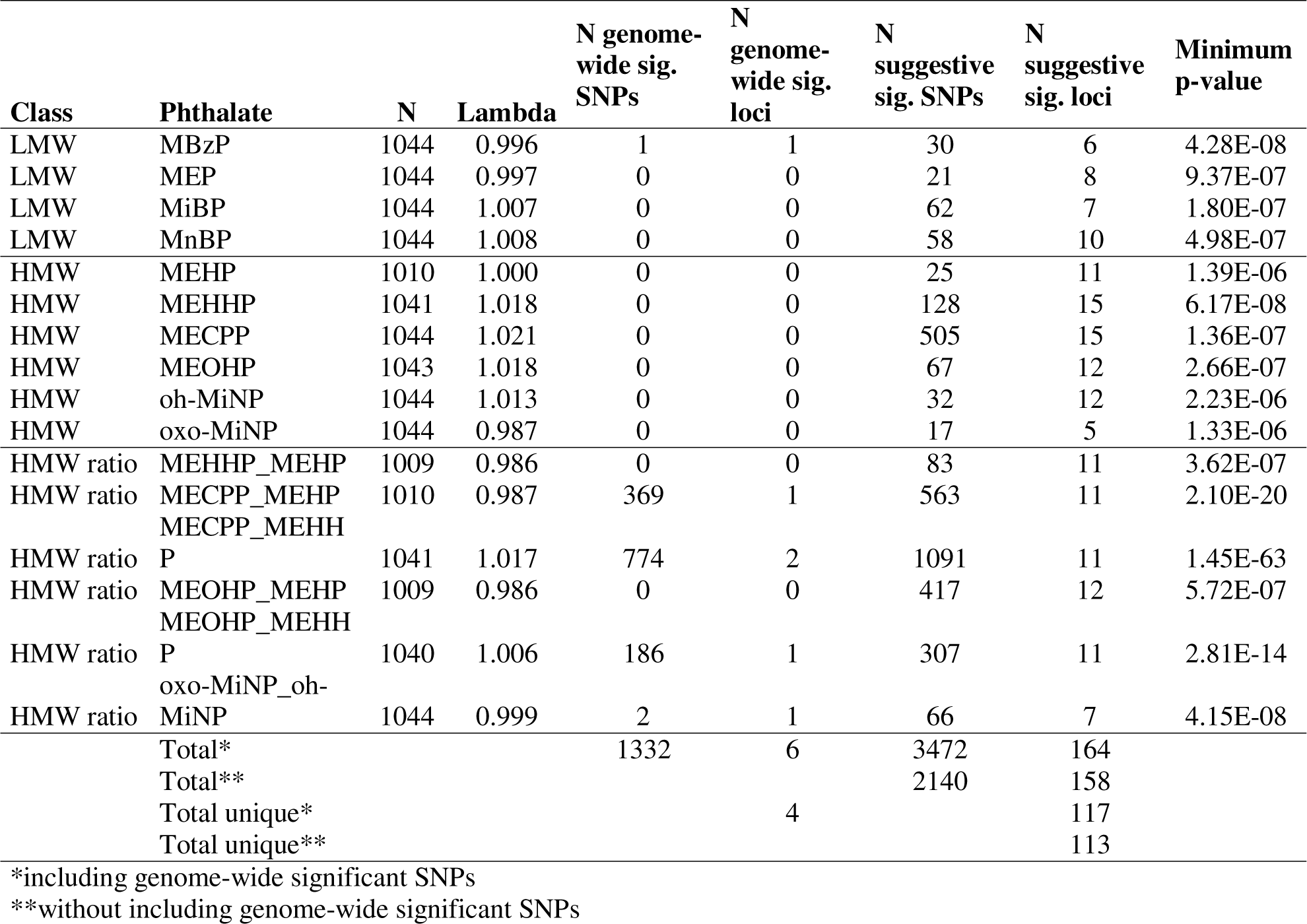
Summary of the results of the single-trait GWAS of phthalate levels and their ratios. A locus is defined as a region of 1 Mb with SNPs with similar effect size on the trait Suggestive statistical significance is set at p-value <1E-05

**Table 3.**
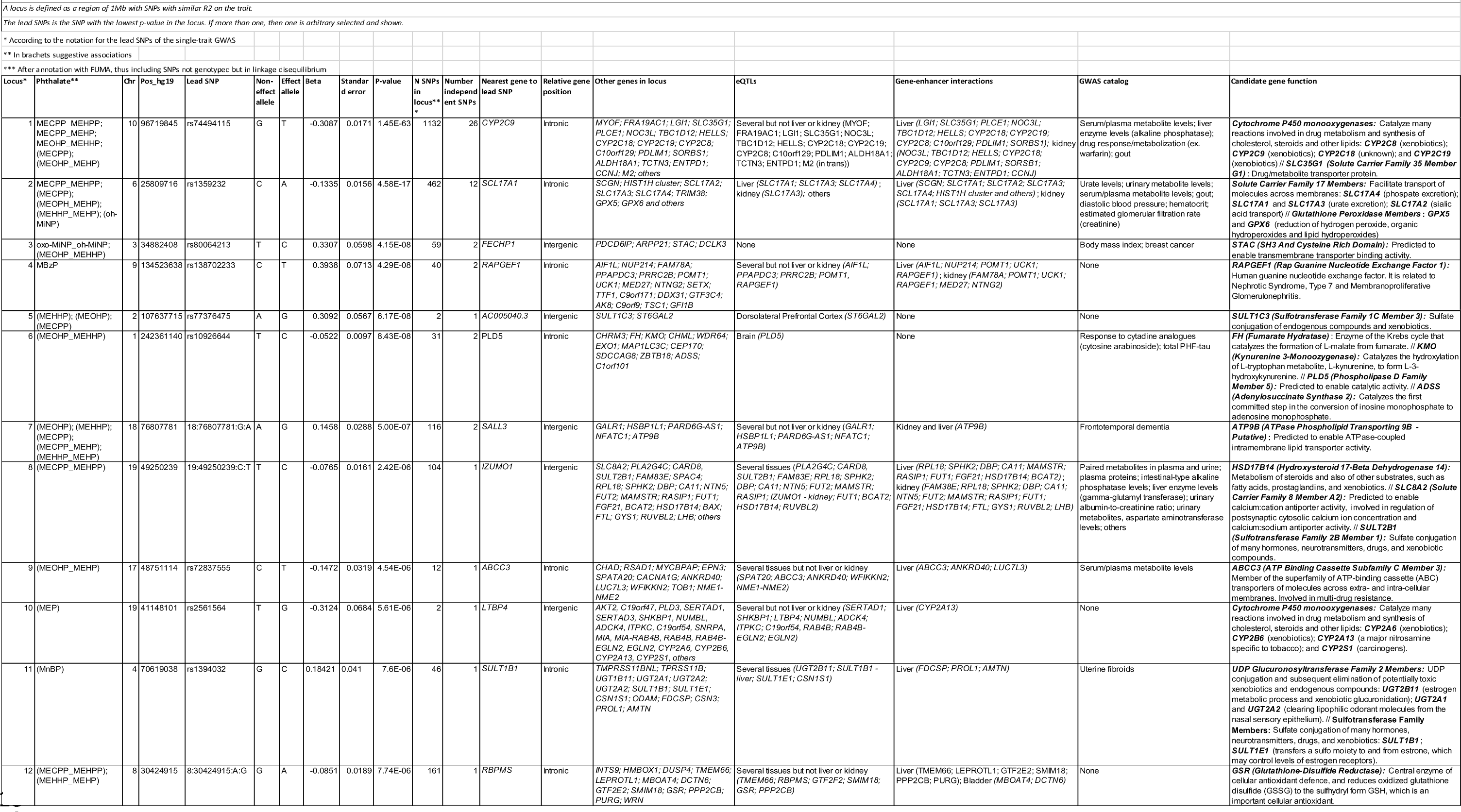
Selected loci associated with phthalate levels and/or ratios, ordered by p-value.

Sensitivity analysis adjusting for creatinine instead that using it to divide phthalate metabolite levels gave similar results with minor differences (Appendix B - Supplementary Table B2). Three of the four genome-wide significant loci had still a p-value <1E-08, and the other hit (chr9 loci associated with MBzP) was marginally significant (p-value = 9.95E-07). A plot comparing p-values and effect sizes between the two models for all the suggestive SNPs can be seen at Appendix A – Supplementary Figure 5A and 5B. Moreover, the GWAS of creatinine levels did not retrieve any genome-wide significant SNP. Only two of the creatinine suggestive SNPs were also suggestively associated with MEOHP and oh-MiNP (Appendinx B Supplementary Table B19).

### 3.3. GWAS of CNVs

Using the PennCNV tool, we identified a median of seven (interquartile range (IQR): 6-9) CNV regions per child with a median length of 43 (IQR: 29-64) kb. Only two children did not present any CNV event. After applying a reciprocal overlap of 50%, 2,916 distinct CNV regions were detected, including 1,785 that were unique, meaning that they were detected in only one child. Thirty-six of the 2,916 CNVs were identified in > 3% of the children and thus labeled as consensus CNV regions and tested in relation to phthalate levels and ratios (Appendix C – Supplementary Table C0).

In 12 of the 36 consensus regions, we detected single or double deletions (loss CNVs, CN=0 and CN=1), in seven of them we detected single duplications (gain CNVs, CN=3) and in 17 of them we detected single and double deletions as well as single duplications (loss and gain CNVs, CN=0, CN=1 and CN=3). We did not detect double duplications (CN=4) in any CNV consensus region. Different models were tested in each type of region (see 2.6.) and the model with the highest significance was reported for each region. The results of the association of these common CNVs with phthalate levels and ratios can be found in Appendix C – Supplementary Tables C3-18.

Fifty-one CNVs presented nominal significance and two of them passed multiple-testing correction (Figure 2, Appendix C - Supplementary Table C1). First, a single-copy gain in CNV region 11A was associated with higher oh-MiNP levels (chr11:18.95Mb-18.96Mb_gain, effect = 1.231, p-value = 1.77E-06, *MRGPRX1* gene), and second, a single-copy gain in CNV region 11B was associated with higher MEP levels (chr11:107.65Mb-107.67Mb_gain, effect = 0.792, p-value = 1.36E-03, *SLC35F2*). The locus zoom plots of these two CNVs are shown in Figure 4.

**Figure 4.**
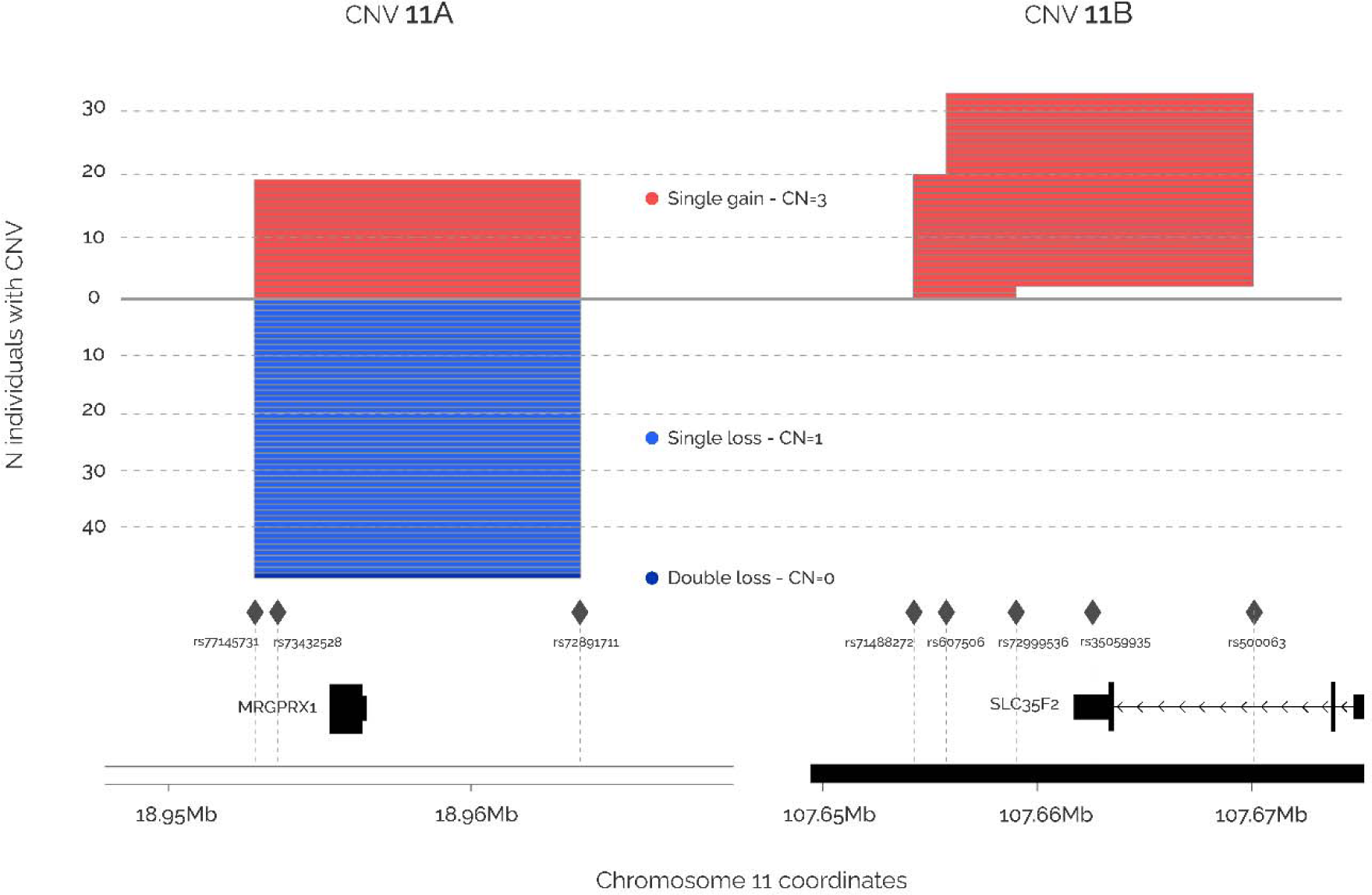
Locus zoom of the consensus CNV-associated regions identified after multiple-testing correction. Each line represents an CNV region in a single sample. Single gain CNVs are coloured in red, single loss CNVs in blue and double loss CNVs in dark blue. The y-axis shows the number of individuals presenting a CNV for the specific region. The x-axis shows the genomic coordinates. Rhombus represent the SNPs used to detect the CNVs, and genes in the regions are also shown.

In the sensitivity analysis adjusting for creatinine, the 88% of the nominally significant associations were reproduced (45 of 51). The associations between CNV 11A and oh-MiNP and between CNV 11B and MEP were very similar (Appendix C - Supplementary Table C2), however the later did not reach the multiple-testing threshold (chr11:18.95Mb-18.96Mb_gain, effect = 1.120, p-value = 2.51E-06; chr11:107.65Mb-107.67Mb_gain, effect = 0.695, p-value =1.48E-03). A plot comparing p-values and effect sizes between the two models for all the suggestive CNVs can be seen at Appendix A – Supplementary Figure 5C and 5D. Results for creatinine GWAS did not show any significant association, with only four CNV regions, not including 11A and 11B, showing a p-value <0.05 (Appendix C - Supplementary Table C19).

### 3.4. Functional annotation of genetic variants and identification of candidate genes and pathways

Next, SNPs with suggestive significance were explored with the FUMA tool providing information on SNP annotation (Appendix D), identification of eQTLs (Appendix E), and comparison with SNPs in the GWAS catalog (Appendix F). Appendix G shows the list of candidate genes in each locus based on FUMA results and gene-enhancer interactions from the EPIraction tool. A selection of 12 interesting loci in terms of statistical significance, annotated genes and/or overlap with GWAS traits is listed in Table 3. These loci are annotated to phase I and phase II detoxification genes [*CYP monooxygenases* (*CYP2C* cluster), *Sulfotransferases* (*SULT1B1*, *SULT1E1*, and *SULT1C3)*, *UDP glucuronosyltransferases* (*UGT2A1*, *UGT2A2* and *UGT2B11*), and glutathione related genes (*GSR*, *GPX5* and *GPX6*)]; and genes related to molecule transport across membranes [*Solute carriers* (*SLC17A* cluster, *SLC8A2*, and *SLC35G1*) and others such as *ABCC3*, *ATP9B* and *STAC*].

After the FUMA annotation, we took suggestive prioritized genes to run gene-set enrichment analyses (Appendix H). Among others, we identified gene-sets related to xenobiotic detoxification, drug detoxification, CYP, oxidoreductase reactions, liver, phase I and phase II enzymes. These gene-sets were found basically for MEP, MECPP and its ratios, and MEOHP ratios.

Finally, we searched for enriched GTEXv8 tissues (Appendix I). At adjusted and/or nominal statistical significance, we found enrichment for candidate tissues involved in phthalate metabolism: liver (MEOHP_MEHP and MEOPH_MEHHP), kidney cortex (MBzP, MECPP_MEHP and MEOHP_MEHHP), and bladder (MECPP_MEHP). Other significant enrichments were found for skin and vagina (oxo-MiNP_oh-MiNP), artery aorta (MiBP), and nerve tibial (MEHP).

## 4. Discussion

In this study we conducted genome-wide screenings of SNPs and CNVs in order to identify genetic variation associated with phthalate metabolism in children. We identified four genome-wide significant loci at chr3 (*FECHP1 –* oxo-MiNP_oh-MiNP), chr6 (*SLC17A1 –* MECPP_MEHPP), chr9 (*RAPGEF1 –* MBzP) and chr10 (*CYP2C9* – MECPP_MEHPP) and two significant CNVs located at chr11 (*MRGPRX1* – oh-MiNP and *SLC35F2 –* MEP). Functional annotation of suggestive SNPs highlighted pathways related to xenobiotic and drug detoxification. Also some of the genes annotated to the SNPs associated with phthalates showed significant enrichment for key tissues as liver and kidney, which is indicative of high gene expression levels in these tissues. Furthermore, previous GWAS have reported significant associations of SNPs located in these loci with drug biotransformation and urinary metabolite levels of several compounds. Below we describe in detail the most relevant loci and genes identified, which are related to phase I and II metabolism, transport of molecules across membranes and kidney function.

### 4.1. Phase I metabolism

We identified three loci annotated to *CYP* genes, one of them achieving the smallest p-value of the study. This top locus at chr10 was associated with four DEHP metabolite ratios (MECPP_MEHPP; MECPP_MEHP; MEOHP_MEHHP; MEOHP_MEHP) and contains *CYP2C8*, *CYP2C9*, *CYP2C18*, and *CYP2C19* genes. According to the EPIraction tool, the locus can act as an enhancer of these genes in liver and/or kidney. *CYP* genes code for phase I monooxygenases that are key in multitude of metabolic processes of endogenous and exogenous compounds, and, indeed, SNPs in this locus have previously been reported to be associated with serum or plasma levels of several metabolites or proteins (Feofanova et al., 2020; Pazoki et al., 2021; Schlosser et al., 2020) and drug response (ie. warfarin) (Takeuchi et al., 2009). Moreover, CYP2C enzymes are known to participate in phthalate metabolism, and *CYP2C9* variants have been associated with the biotransformation of MEHP into secondary metabolites in young adults (Stajnko et al., 2022). Our study confirms that similar detoxification mechanisms take place in children. The two additional *CYP* loci, one at chr7 harboring *CYP2W1* and the other at chr19 including *CYP2A6*, *CYP2B6*, *CYP2A13* and *CYP2S1*, showed suggestive associations with urinary MEP levels (metabolite of diethyl phthalate (DEP)). Therefore, according to our results, DEP (LMW phthalate present in many personal care products, particularly those containing fragrances) and DEHP (HMW phthalate mainly used for food packaging) seem to be metabolized through different CYP genes.

### 4.2. Phase II metabolism

We also identified several loci annotated to phase II metabolism genes, including *UDP*LJ*glucuronosyltransferases* (*UGTs*), *Sulfotransferases* (*SULTs*) and *Glutathione S*LJ*transferases* (*GSTs*). First, we found a locus at chr4, annotated to *UGT2B11*, *UGT2A1* and *UGT2A2,* to be suggestively associated with levels of MnBP, a metabolite of di-n-butyl-phthalate (DnBP), which is found in paints, adhesives, personal care products and deodorants. SNPs in another family member, *UGT1A7*, were previously related to urinary levels of 2-cyclohexane dicarboxylic acid, diisononyl ester (DINCH) metabolite in men and of diisobutyl phthalate (DiBP) and dibenzyl phthalate (DBzP) metabolites in women (Stajnko et al., 2022). Notably, DiBP and DnBP are structural isomers. Second, we found three loci annotated to *Sulfotransferases*: the same locus at chr4 associated with MnBP also contained the *SULT1B1* and *SULT1E1* genes, and two additional suggestive loci at chr2 and chr19 harboring SNPs near *SULT1C3* and *SULT2B1* genes, respectively, that showed associations with DEHP metabolites. Previous studies have described that multiple phthalates have strong inhibition potential towards *SULT* enzymes, potentially causing reduced body detoxification and liver injury (Ceauranu et al., 2023; Huang et al., 2022). Finally, we identified two loci that contain genes that participate in the glutathione metabolism: a genome-wide significant locus at chr6 annotated to *GPX5*, *GPX6* and *SLC* genes associated with MEHP and oh-MiNP levels; and a suggestive locus at chr8 near *GSR*, a central enzyme of cellular antioxidant defence, associated with MEHP ratios. This is consistent with previous research reporting that in zebrafish, MEHP exposure alters the expression of *gsr* (Kwan et al., 2021).

### 4.3. Molecular transport across membranes and kidney function

Besides detoxification, we identified several loci annotated to transmembrane transporter genes which could potentially play a role in the excretion of phthalates in the intestine or kidney. These encompass several *Solute carrier* (*SLC*) genes that code for transporters of a wide array of substrates across membranes (Liu, 2019). A locus at chr6 harboring *SLC17A1*, *SLC17A2*, *SLC17A3* and *SLC17A4* was associated with MEHP ratios at genome-wide significance and showed suggestive associations with oh-MiNP. Both gene-enhancer and eQTL information, suggested that these SNPs regulate *SLC* expression in liver and/or kidney. Moreover, it is known that *SLC17A1* and *SLC17A3* participate in urate regulation by facilitating the excretion of intracellular urate from the bloodstream into renal tubule cells. Consistently, previous GWAS reported associations between this locus and serum uric acid levels (Hollis-Moffatt et al., 2012), urinary metabolite levels (Schlosser et al., 2020), estimated glomerular filtration rate (creatinine) (Stanzick et al., 2021), and chronic kidney disease (Torres et al., 2021), among others. Besides, other associations involving the *SLC* family were identified, including the association between a single-copy gain in a CNV that overlaps the final exon and 3’UTR of the *SLC35F2* gene, predicted to enable transmembrane transporter activity, and MEP urinary levels.

*ABCC3* encodes a member of the ATP-binding cassette (ABC) transporters that play a role in the transport of biliary and intestinal excretion of organic anions. Moreover, it is reported to be involved in multi-drug resistance (Deeley et al., 2006; Ramírez-Cosmes et al., 2021) and variants in the gene are related to serum/plasma metabolite levels (Bruhn and Cascorbi, 2014). In our study, SNPs in the *ABCC3* intronic region (chr17) were found to be suggestively associated with the MEOHP_MEHP ratio. Notably, another study showed that flies exposed to dibutyl-phthalate (DBP) have increased expression of several genes, including an homologous of the *ABCC3* gene (Williams et al., 2016). Two more transporters were identified. On the one hand, SNPs in a liver and kidney enhancer for *ATP9B* gene (chr18), which is predicted to enable ATPase-coupled intramembrane lipid transport, were suggestively associated with DEHP metabolites and ratios. On the other hand, SNPs near *STAC* (chr3) were associated with the oxo-MiNP_oh-MiNP ratio at genome wide significance and with the MEOHP_MEHHP ratio at suggestive significance. *STAC* is predicted to enable transmembrane transporter binding activity and participate in positive regulation of voltage-gated calcium channels and skeletal muscle contraction.

The last group of results is comprised by two genes potentially implicated in kidney function: *RAPGEF1* and *MRGPRX1*. First, SNPs located in a liver and kidney enhancer for *RAPGEF1* at chr9 were genome-wide associated with MBzP, a metabolite of butylbenzyl phthalate (BBP) used as plasticizer for polyvinyl chloride (PVC). *RAPGEF1* encodes a guanine nucleotide exchange factor, and mutations in genes of the same family have been found in patients with nephrotic syndrome type 7 and membranoproliferative glomerulonephritis (Maywald et al., 2022; Zhu et al., 2019). Second, a loss-gain CNV of 10.8 kb overlapping the whole gene *MRGPRX1* was associated with oh-MiNP, a metabolite of DINP which is used primarily in the manufacture of PVC. *MRGPRX1* encodes a Mas-related G-protein-coupled receptor reported to be responsible for itch sensation, pain transmission and inflammatory reactions, including those to the antimalarial drug chloroquine (Gan et al., 2023). Additionally, mutations in other G-protein-coupled receptors have been implicated in nephrogenic diabetes insipidus, a disease characterized by polyuria and polydipsia (Wang et al., 2018). We found that individuals with three copies of the gene had increased levels of urinary levels of oh-MiNP, whereas individuals with zero or one copy had no difference versus the reference group with two copies. Given the low numbers presenting the alternative allele, further investigation in a larger sample would be required.

Our study found several associations between phthalates and SNPs known to regulate urinary metabolite levels, glomerular filtration rate, and kidney function. These associations might indicate a real participation of these SNPs in phthalate excretion. Nevertheless, as our phthalate measurements were normalized by creatinine to account for urine dilution, some of our results could also just reflect renal excretion capacity. Although both explanations are possible, there is some evidence supporting the former. First, in vitro studies suggest that phthalates can be a substrate for the *SLC* transporters (Klaassen and Aleksunes, 2010); second, the associations found with the phthalate ratios are unaffected by the creatinine normalization; third the sensitivity model adjusting for creatinine instead of normalizing by it provided similar results; and four the GWAS of creatinine did not found many suggestive loci overlapping with loci found for phthalates.

### 4.4. Strengths of the study

The study has several strengths. First, we measured a large number of phthalate metabolites using a robust analytical method in an equal volume daily pool of two urine samples (morning and night). Second, instead of analyzing individual SNPs in candidate genes, we performed genome-wide screenings of SNPs and CNVs and several downstream annotation methods that allowed us to identify novel loci implicated in phthalate metabolism. Although some genes seemed to be involved in detoxification of more than one phthalate compound (ie. *SULT1C3* for MnBP and MEHP, or *SLC17* for oh-MiNP and MEHP), others appeared to be more specific (ie. *CYP2C* for MEHP while *CYP2A* for MEP). Third, the study was conducted in a large sample of children from the HELIX project, higher than previous similar studies but still limited in comparison with GWAS of other traits. This study, besides producing novel knowledge about the genes participating in phthalate detoxification and renal excretion, also provides a list of SNPs that can be used to stratify individuals in epidemiological research. Bearing the “alternative” alleles of these SNPs can confer different susceptibility to phthalate’s effects depending on the SNP function (i.e., fast biotransformation to a neutral compound and then excretion versus fast biotransformation to a more toxic compound).

### 4.5. Limitations of the study

However, the study also has some limitations. First, we have restricted our discussion on few genes with interesting functions for phthalate metabolism. However, since SNPs can regulate long distance genes, other genes not highlighted here could also be relevant. Functional analyses in in vitro studies or animal models should be conducted in order to identify the causal variants and genes. Second, we analyzed phthalate metabolites in a pool of two urines and then standardized for creatinine. This has some limitations, especially for pooled samples as discussed elsewhere (O’Brien et al., 2017; Philippat and Calafat, 2021), but currently there is no consensus on how to account for urine dilution in equal volume pools. As discussed above, this could have resulted in the identification of genetic variants that regulate glomerular filtration in general, rather than phthalate excretion in particular. Finally, the analyses were restricted to European ancestry children, thus the translation to other ancestries is unknown.

## 5. Conclusions

In summary, through genome-wide screenings we identified known and novel loci potentially implicated in phthalate metabolism in children. Genes annotated to these loci participate in phase I and phase II metabolism and renal excretion process. Replication of the findings in independent studies and validation in in vitro systems is required.

## Data Availability

The raw data supporting the current study are available from the corresponding author on request subject to ethical and legislative review. The "HELIX Data External Data Request Procedures" are available with the data inventory in this website: http://www.projecthelix.eu/data-inventory.

## Abbreviations

ABC: ATP-binding cassette
BBP: butylbenzyl phthalate
chr: chromosome
CNV: copy number variant
CYP: Cytochrome P450 monooxygenase
DBzP: dibenzyl phthalate
DEP: diethyl phthalate
DEHP: bis(2-ethylhexyl) phthalate
DiBP: diisobutyl phthalate
DINCH: 2-cyclohexane dicarboxylic acid, diisononyl ester
DnBP: di-n-butyl-phthalate
EDCs: endocrine disrupting chemicals
GSA: Infinium Global Screening Array
GWAS: genome-wide association study
HELIX: Human Early Life Exposome project
HMW: high-molecular-weight
HRC: haplotype reference consortium
LD: linkage disequilibrium
LMW: low-molecular-weight
MBzP: mono benzyl phthalate
MECPP: mono-2-ethyl-5-carboxypentyl phthalate
MECPP_MEHP: ratio between MECPP and MEHP
MECPP_MEHHP: ratio between MECPP and MEHHP
MEHHP: mono-2-ethyl-5-hydroxyhexyl phthalate
MEHHP_MEHP: ratio between MEHHP and MEHP
MEHP: mono-2-ethylhexyl phthalate
MEOHP: mono-2-ethyl-5-oxohexyl phthalate
MEOHP_MEHP: ratio between MEOHP and MEHP
MEOPP_MEHHP: ratio between MEOPP and MEHHP
MEP: monoethyl phthalate
MiBP: mono-iso-butyl phthalate
MnBP: mono-n-butyl phthalate
oh-MiNP: mono-4-methyl-7-hydroxyoctyl phthalate
oxo-MiNP: mono-4-methyl-7-oxooctyl phthalate
oxo-MiNP_oh-MiNP: ratio between oxo-MiNP and oh-MiNP
PVC: polyvinyl chloride
QQ: quantile-quantile
SCL: Solute carrier
SNP: single nucleotide polymorphism
SULT: Sulfotransferase
UGT: UDP-glucuronosyltransferase

## 7. Author Contributions

MB conceptualized the study. MB, LB, ZB, and RN conducted the statistical analysis or functional enrichment analysis with the support of JRG. The HELIX project was coordinated by MV with the support of LM. SA, ALB, MAC, MC, LC, RG, KBG, BH, JL, JS, TCY, JW, are the PIs of the cohorts or participated in sample or data acquisition. AKS was responsible for measurement of phthalate metabolites. MB, GE and CRA participated in genetic data acquisition. LB, ZB and MB wrote the original draft of the paper and all the other co-authors contributed to reviewing and editing the manuscript.

## 8. Acknowledgements

We would like to thank all the families for their generous contribution.

## 10. Data Availability Statement

Summarized results can be found at https://helixomics.isglobal.org/ **(THIS WILL BE DONE UPON ACCEPTANCE OF THE MANUSCRIPT).** The raw data supporting the current study are available from the corresponding author on request subject to ethical and legislative review. The “HELIX Data External Data Request Procedures” are available with the data inventory in this website: http://www.projecthelix.eu/data-inventory.

