## Appendix A for "Common genetic variants associated with urinary phthalate levels in children: a genome-wide study"

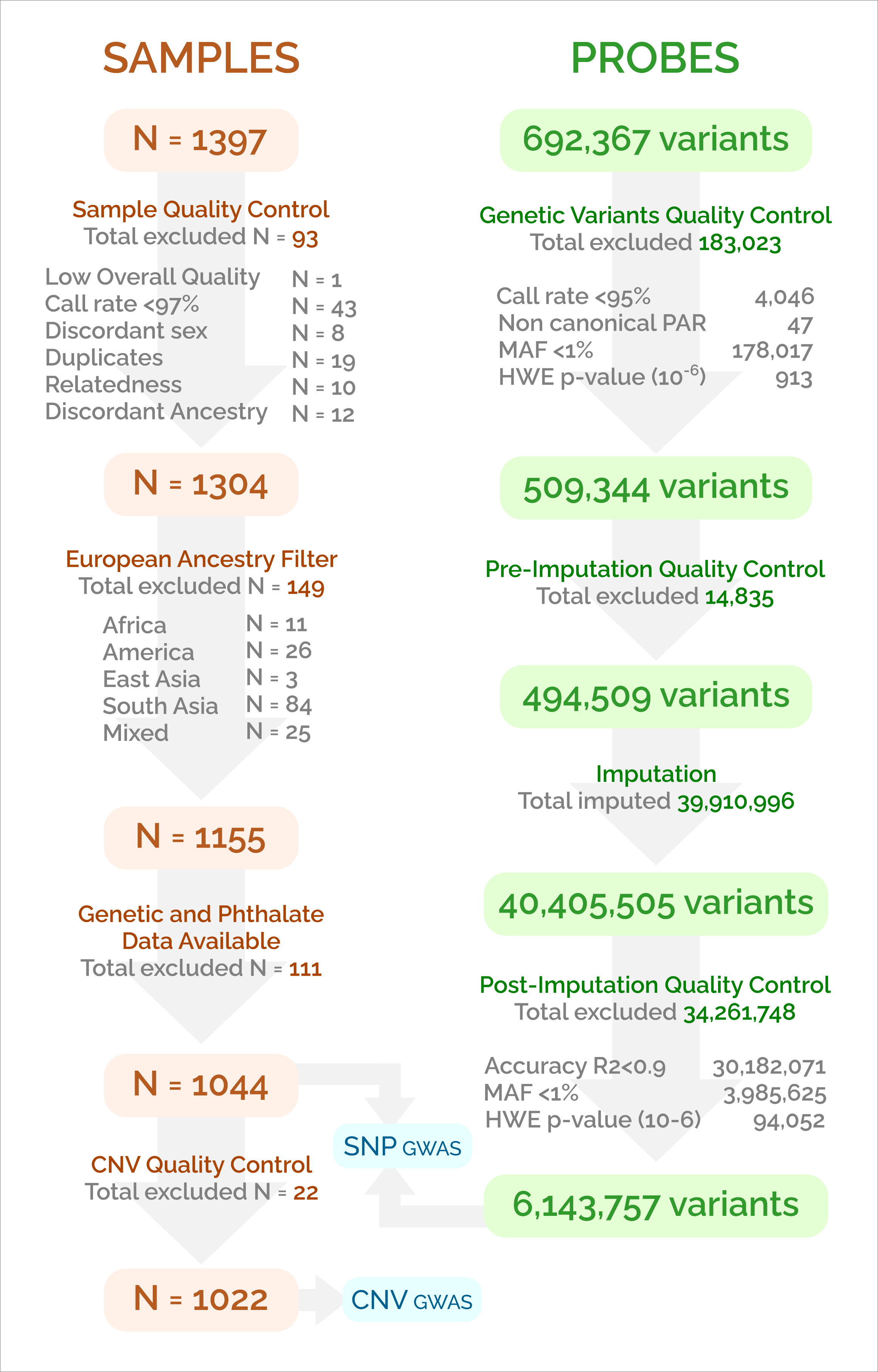

**Appendix A**

**Supplementary Figure 1. Flow chart of the quality control of samples and probes**

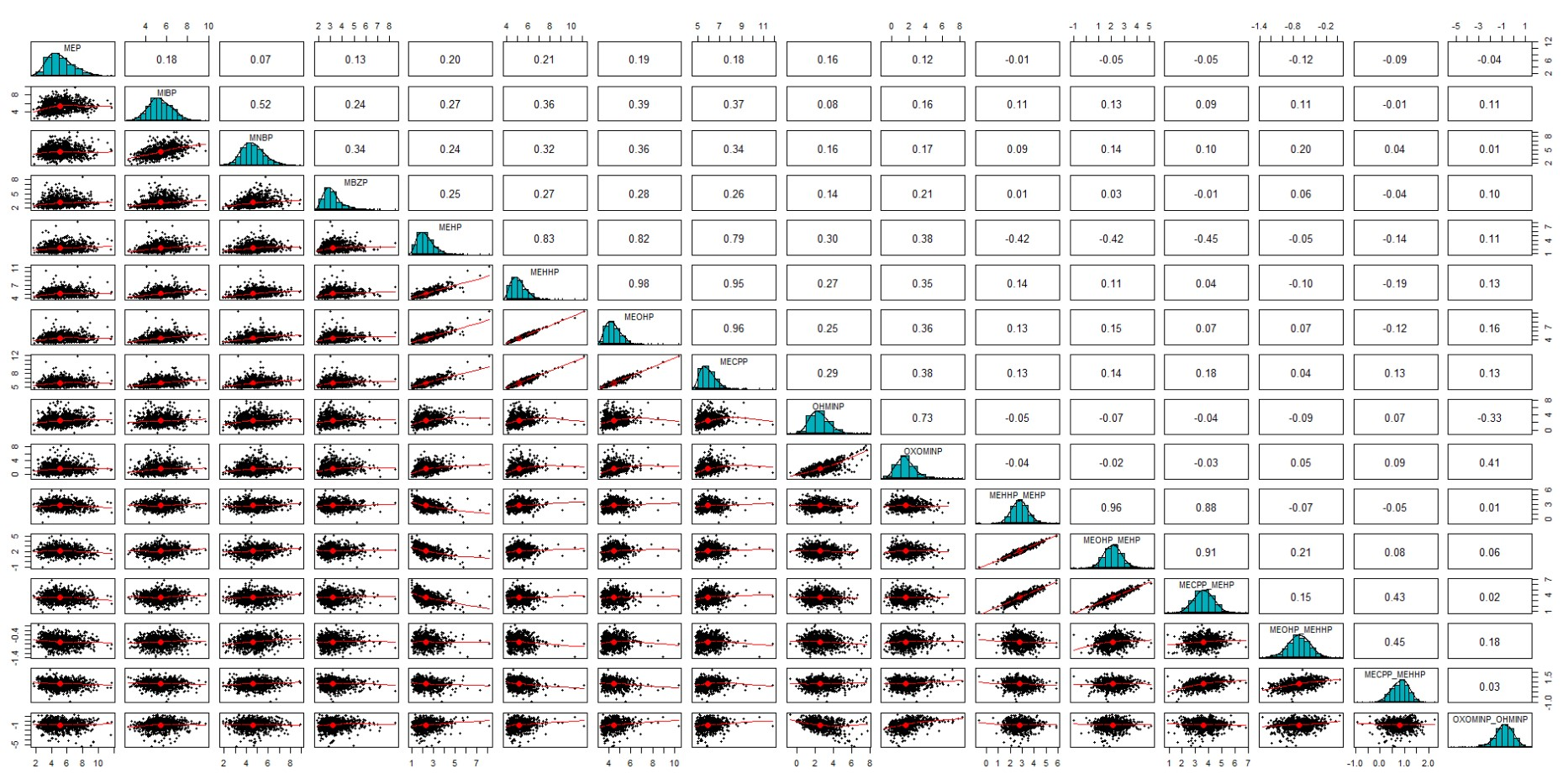

**Supplementary Figure 2. Pair-wise correlation between phthalates, ordered alphabetically**

The x- and y-axis represent the log2 phthalate levels. The pairwise-correlation is shown in the upper panels, while the correlation plot in the lower panels. The diagonal panels show the distribution of phthalate levels.

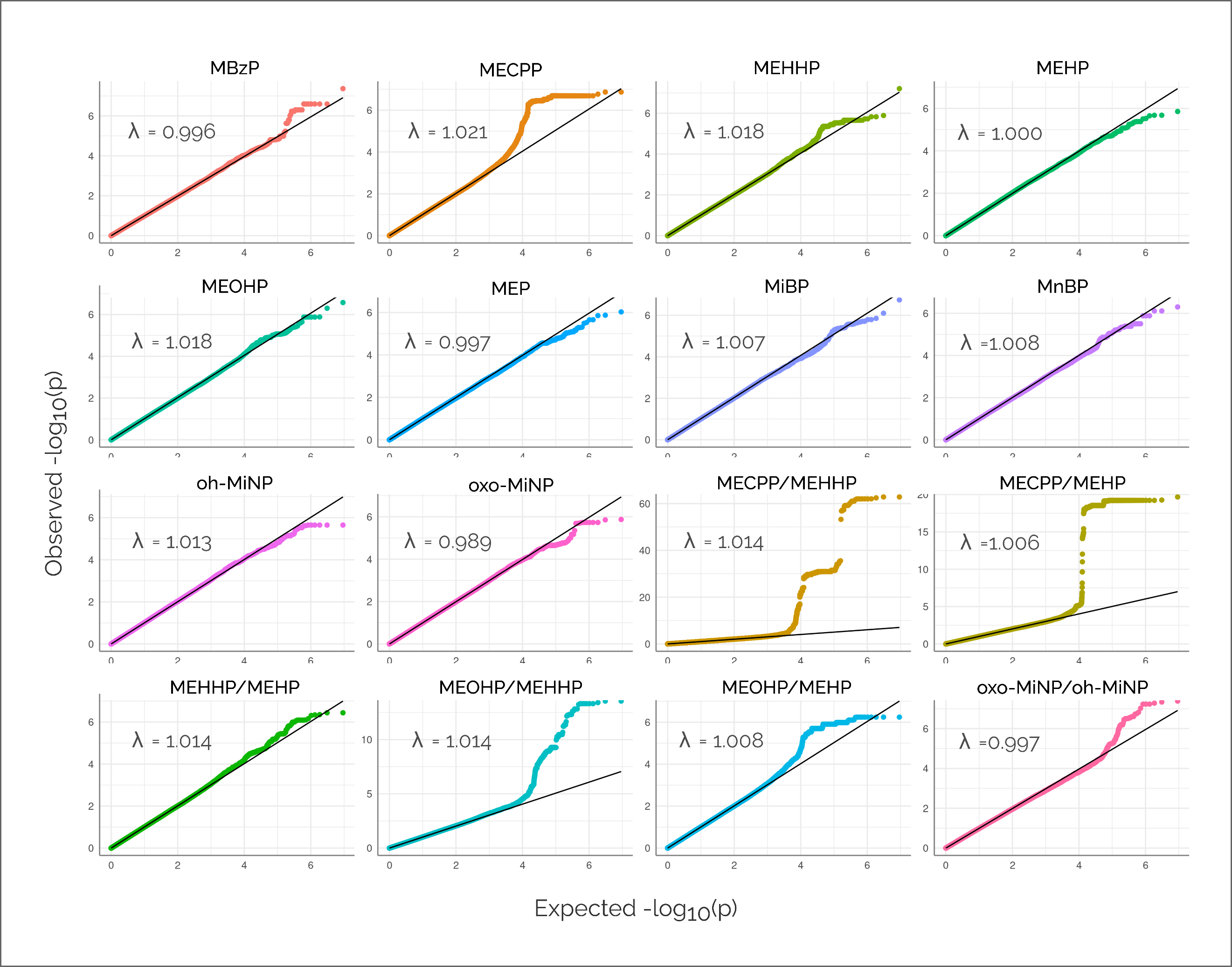
**Supplementary Figure 3. QQ plots for all phthalates and ratios from single-trait GWAS**

The x-axis represents the expected –log10 p-value under the null hypothesis and the y-axis the observed –log10 p-value of the association between genome-wide genetic variants and phthalate levels or ratios.

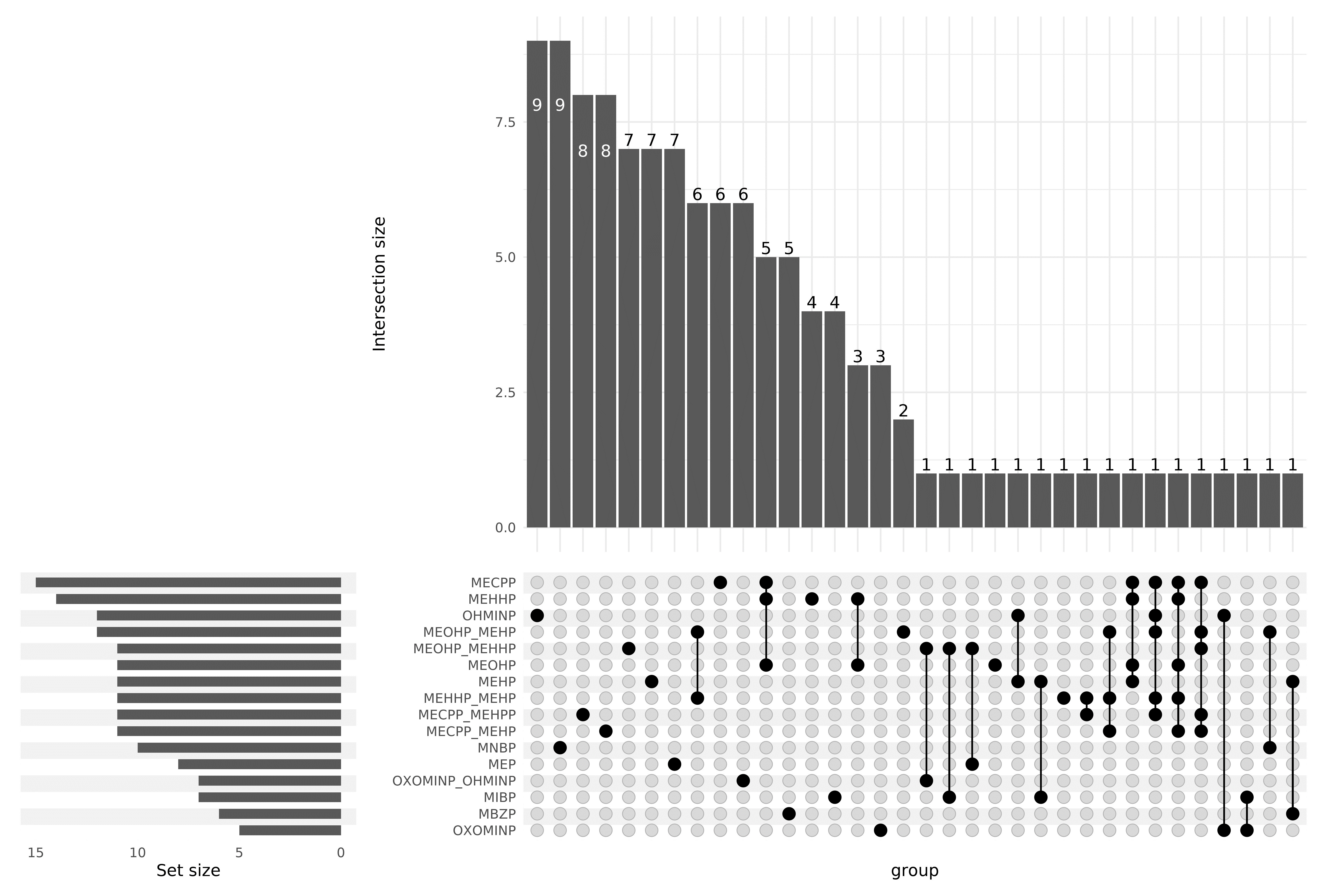

**Supplementary Figure 4. Overlap of suggestive loci (p-value <1E-05) across phthalates and ratios from the single-trait GWAS**

Set size indicates the number of loci identified for each phthalate and ratio. Intersection size indicates the number of loci which are common across phthalates and/or ratios.

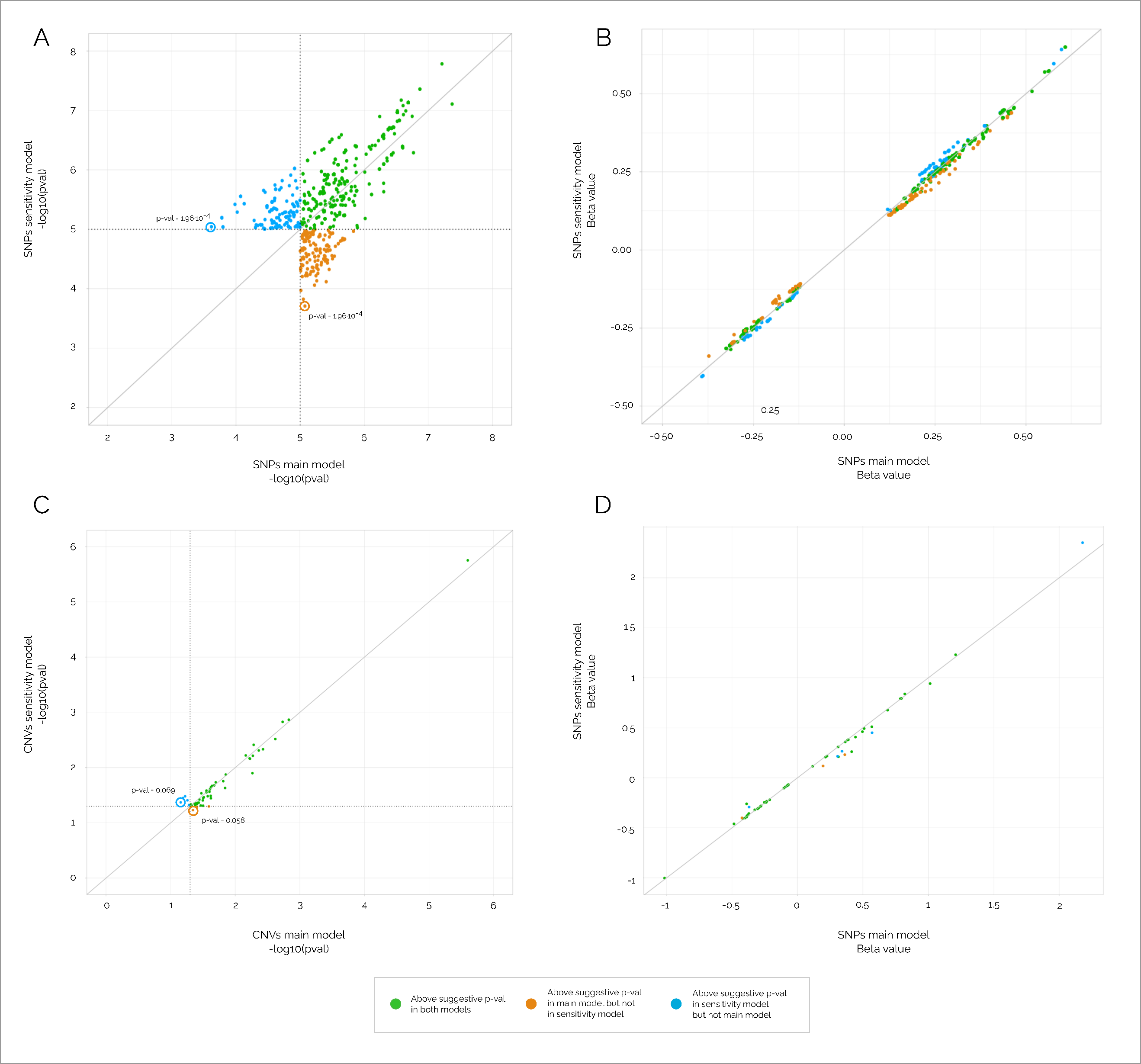

**Supplementary Figure 5. Comparison of the results of the main model (phthalate/creatinine ~ genotype + sex + age + 10 GWAS PCs) vs the results of the sensitivity model (phthalate ~ genotype + sex + age + 10 GWAS PCs + log2(creatinine))**

For SNP GWAS, panel A shows the correlation of p-values and panel B the correlation of effect sizes. For CNV GWAS, panel C shows the correlation of p-values and panel D the correlation of effect sizes. X-axis represents the results of the main model and y-axis the results of the sensitivity model. Each dot is an association and colours indicate whether the association was detected at suggestive p-value in both models (green), in only the main model (orange), or only in the sensitivity model (blue). Associations with the highest p-value among the suggestive ones are indicated with a circle.
